# Summary Estimates Derived from a Multi-state Non-Markov Framework to Characterize the Course of Heart Disease

**DOI:** 10.1101/2024.09.18.24313882

**Authors:** Ming Ding, Feng-Chang Lin, Michelle L. Meyer

## Abstract

Multi-state Markov models have been used to model the course of chronic disease. However, they are unsuitable to chronic disease where past and present states interplay and affect future states, and the estimated transition probabilities are time-specific which are not straightforward for public health interpretation. We have proposed a multi-state non-Markov framework that splits disease states into substates conditioning on past states. As the substates track past states and indicate multimorbidity, the estimated transition rates can be used to derive two summary estimates: Disease path which shows path of state transition, and multimorbidity-adjusted life year (MALY) which represents the adjusted life year in full health. In this paper, we showed the derivation of the two summary estimates and applied them to characterize the course of heart disease using data from the Atherosclerosis Risk in Communities Study (ARIC) study. The course of heart disease was modeled in five states, namely, healthy, at metabolic risk, coronary heart disease (CHD), heart failure, and mortality. In this mid- to old-age population, the estimated MALY was 24.13 (95% CI: 16.55, 32.06) years. For healthy participants at baseline, the most likely disease paths were: “Healthy → at metabolic risk → mortality” (37%), “Healthy → mortality” (21%), “Healthy → at metabolic risk → heart failure → mortality” (19%), and “Healthy → at metabolic risk → CHD → mortality” (8%). The MALY was higher among women than men and higher among Whites than Blacks. The distribution of disease path was similar across sex and race subgroups. In summary, MALY and disease path characterize the disease course in a summary manner and have potential use in chronic disease prevention.

## 1. INTRODUCTION

Multi-state models, which fit one survival model to each state, have been used to describe course of chronic disease. Most of the existing methods are Markov models, which assume that present and future states are independent of past history.^1, 2^ Markov models are unsuitable to describe the course of chronic disease for several reasons. First, for chronic disease, past and present states interplay and affect future states which violates the Markov assumption. Second, by assuming present state is independent of past history, Markov models do not allow for multimorbidity, a common condition for chronic disease.^3–5^ Finally, the transition probabilities estimated from Markov models are time-specific, and these high-dimensional matrices are not straightforward for public health interpretation.

Non-Markov models are a largely unexplored field, with only several non-parametric methods proposed,^6–12^ such as the landmark Aalen-Johansen (LMAJ) estimator.^6^ We have proposed a multi-state non-Markov regression framework to investigate the course of chronic disease.^13^ By conditioning on past disease history, disease states are divided into substates to convert non-Markov to Markov process, where cause-specific Cox models are used to estimate transition rate between substates. As substates memorize disease history and track multimorbidity, the transition rates between substates can be synthesized into summary estimates to characterize the whole disease course: Disease path, and multimorbidity-adjusted life year (MALY) which represents the adjusted life year in full health. The two summary estimates are easy to interpret and can be used for Markov process (as a special case for non-Markov process), which may advance multi-state modeling in disease prevention. In this paper, we showed the derivation of the two summary estimates from our non-Markov framework and applied them to characterize the course of heart disease in a large cohort study.

## 2. METHODS

### 2.1. An introduction to the multi-state non-Markov framework

We developed a non-Markov regression framework that allows past states to affect transition rates of current states.^13^ While it can be used for chronic disease of any states, we assume five states S_0_-S_4_ for illustration purpose (**Figure 1a**). We assume that transition occurs between any two states, while our method works if there is no transition between two states. The details of our framework are shown below.

**Figure 1.**
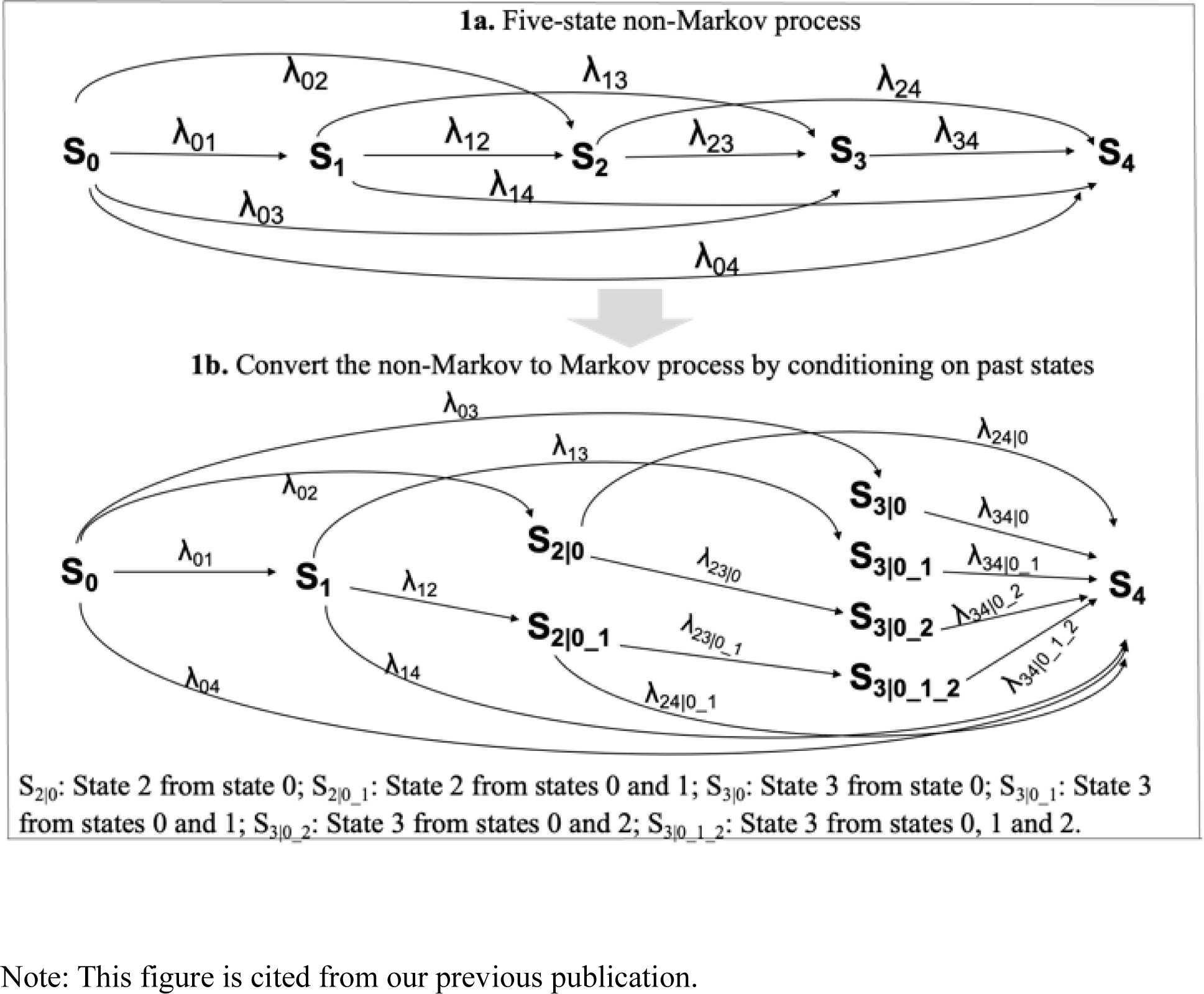
Convert non-Markov to Markov process by splitting disease states into substates.^13^.

#### Convert a non-Markov to Markov process

Our framework allows for non-Markov process, where past disease states can affect current and future states. However, the Aalen-Johansen (A-J) estimator is systematically biased in non-Markov process, causal difficulty in estimation of transition probabilities.^10^ Thus, we divide disease states into substates by conditioning on past disease history to convert the non-Markov process to Markov process (**Figure 1b**). As the substates are independent of past disease states, the converted process in **Figure 1b** satisfies the Markov assumption.^13^

#### Construct transition rate matrix between substates 𝐐(𝐭)

The matrix of transition rates can be constructed Based on **Figure 1b** (**Figure 2**). Each row and each column stand for a disease substate, and 𝐐(𝐭)[a,b] stands for the transition rate from the substate in row a to the substate in column b. If there is no transition between two substates, the transition rate is 0.

**Figure 2.**
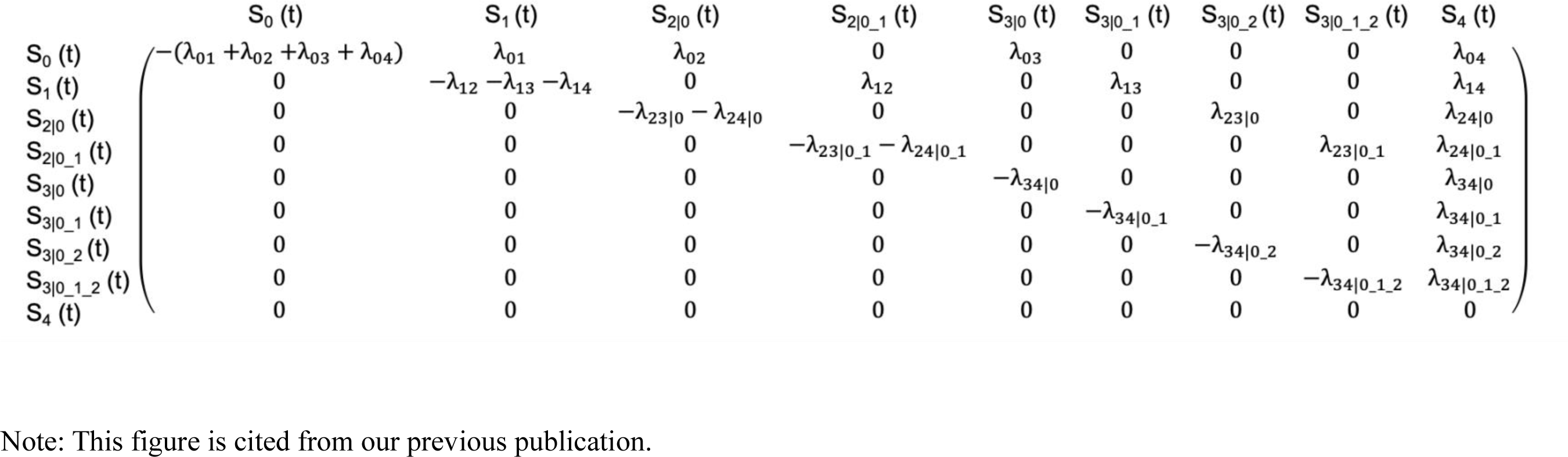
Transition rate matrix Q(t).^13^.

#### Estimate transition rates 𝐐(𝐭)

The transition rate matrix is estimated by fitting multiple cause-specific Cox (CSC) models, which was described in detail in our previous paper.^14^ In brief, we subset the data by disease states and change the data from wide to long structure by age. CSC model is fitted to each subset to estimate transition rates.^13^ As the data is changed into long structure by age, the predicted risk at the end of each time interval is approximate to age-specific hazard in discrete time, which can be estimated using cumulative incidence function (CIF) to account for competing risks.^13^

#### Estimate state occupational probabilities 𝐏(t). 𝐏(t) =

(p_0_(t), p_1_(t), p_2|0_(t), p_2|0_1_(t), p_3|0_(t), p_3|0_1_(t), p_3|0_2_(t), p_3|0_1_2_(t), p_4_(t)), where p(t) is the probability of participants in a particular state. Based on the discrete-time A-J estimator, the change in 𝐏(t) from t to t+1 can be estimated as 𝐏’(t) = 𝐏(t) ∗ 𝐐(t). Thus, 𝐏(t_2_ + 1) = 𝐏(t = t_1_) ∗ 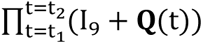, where 𝐐(t) is the transition rate matrix and I_9_ an identity matrix. In this paper, based on the age-specific transition rates 𝐐(𝐭) and state occupational probabilities 𝐏(𝐭), we derive two summary estimates, MALY and disease path.

### 2.2. Estimate MALY

To estimate MALY, we first estimate disease-state specific life year (SSLY), which is calculated as sum of state occupation probabilities in each state from start age until death. Specifically, SSLY in substate k from 𝑡_1_ to 𝑡_2_ can be estimated as 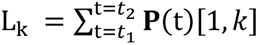, where 𝐏(t)[1, 𝑘] is the k^th^ column of 𝐏(t). The substate k ranges from 1 to 8 refers to substates S_0_, S_1_, S_2|0_, S_2|0_1_, S_3|0_, S_3|0_1_, S_3|0_2_, and S_3|0_1_2_, respectively. We stop estimating 𝐏(t) at age t_2_ if 𝐏(t)[1,9]>0.95, as we assume state occupational probability for death>0.95 indicates a participant’s death.

The substates identified from our non-Markov framework inform multimorbidity. For example, S_3|0_1_ shows a participant in state S_3_ with a history of S_0_ and S_1_ and indicates multimorbidity of S_1_ and S_3_. MALY is calculated as an average of SSLY across substates weighted by multimorbidity, where the weight is calculated using a multiplicative approach.^15, 16^ Particularly, we assign a disability weight (DW) to substates without multimorbidity (i.e., S_0_, S_1_, S_2|0_, S_3|0_), which measures the severity of health state from disease or injury. DW has a value of 0-1, with 0 indicating full health and 1 indicating death.^15^ We use a multiplicative approach to assign DW to states with multimorbidity (i.e., S_2|0_1_, S_3|0_1_, S_3|0_2_, and S_3|0_1_2_).^16^ For example, for multimorbidity with two diseases (e.g., S_2|0_1_), DW_2|0_1_ 1 − (1 − DW_2_)(1 − DW_1_), and for multimorbidity with three diseases (S_3|0_1_2_), DW_3|0_1_2_ 1 − (1 − DW_1_)(1 − DW_2_) (1 − DW_3_). As all substates are mutually exclusive, MALY ∑_𝑘_ DW_k_ ∗ L_k_, which is as a weighted sum of SSLY across substates.

### 2.3. Estimate disease path

Path of disease can be estimated by decomposing death into probabilities through each disease path. We use state occupation probabilities multiplied by transition rates to estimate age-specific probability of death through each path, and then sum up the probabilities over time to obtain probability of each path. By applying 𝐏’(t) = 𝐏(t) ∗ 𝐐(t), change in 𝐏(t + 1) from 𝐏(t) can be calculated for each substate. Particularly, 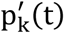, which is change in 𝐏(t + 1) from 𝐏(t) for substate k, can be estimated as 𝐏(t) ∗ (𝐐(t)[, k]). Thus, the probability of increase in state of death from time t to t + 1 is 𝐏(t) ∗ (𝐐(t)[, 9]); the probability of this increase in death transited from substate k is (𝐏(t)[1, k]) ∗ (𝐐(t)[𝑘, 9]); and the probability of death from time 𝑡_1_to 𝑡_2_ transited from substate k are 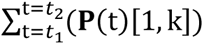 ∗ (𝐐(t)[𝑘, 9]). Specifically, for k 1 to 9, 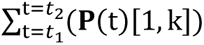 ∗ (𝐐(t)[𝑘, 9]) calculates probabilities of death transited from S_0_, S_0_→S_1_, S_0_→S_2_, S_0_→S_1_→S_2,_ S_0_→S_3_, S_0_→S_1_→S_3,_ S_0_→S_2_→S_3,_ S_0_→S_1_→S_2_→S_3,_ and S_4_, respectively.

#### Use bootstrap to obtain 95% CIs

We adopted a parametric bootstrapping approach to repeatedly draw transition rates from their corresponding distributions to estimate the variability of the parameter estimate. We repeated the resampling process 100 times in each estimation by the CSC model. Statistically, the resulting 100 parameter estimates represent the empirical distribution. MALY are summarized with median values and 95% CI defined by 2.5th to 97.5th percentiles of the empirical distribution, and probabilities of each disease path is calculated over the 100 estimates.

## 3. APPLICATION

### 3.1. Study population

The Atherosclerosis Risk in Communities Study (ARIC) study was designed to investigate the causes of atherosclerosis and its clinical outcomes, as well as variations in cardiovascular risk factors and disease by sex and race.^17^ The enrolled participants ARIC underwent a phone interview and clinic visit at baseline and were followed up by telephone calls and re-examinations from 1987-2019. Participants were contacted periodically by phone and interviewed about interim hospital admissions, cardiovascular outpatient diagnoses, and deaths. Participants who reported CVD-related events were asked to provide medical records that were reviewed by physicians.^18^ For each event, the month and year of diagnosis were recorded as the diagnosis date. Heart failure was ascertained by surveillance calls or clinic visits and was verified from death certificates, medical records, and outpatient diagnoses. Deaths were identified from systematic searches of the vital records of states and of the National Death Index, supplemented by reports from family members and postal authorities.^19^ We obtained ARIC data through Biologic Specimen and Data Repository Information Coordinating Center (BioLINCC), an open repository created by the National Heart, Lung, and Blood Institute (NHLBI), with the Institutional Review Board (IRB) deemed exempt by the University of North Carolina (UNC)-Chapel Hill Review Board.

### 3.2. Methods

We modeled the course of heart disease in five states: Healthy, at metabolic risk, coronary heart disease (CHD), heart failure, and mortality. At metabolic risk state was defined as development of hypertension, hyperlipidemia, or diabetes. Hypertension was defined as blood pressure ≥140/90 mmHg or a history of hypertension or use of blood pressure medications.^20^ Hyperlipidemia included primary hypertriglyceridemia (≥175 mg/dL) or primary hypercholesterolemia (LDL-c 160–189 mg/dL, and/or non-HDL-c 190–219 mg/dL).^21^ Diabetes was defined as fasting glucose ≥7.7mmol/L.^22^ We calculated the time of incidence of at-risk state as the earliest time that any of the risk factors were documented. For participants entering CHD and mortality states at the same time (i.e., incident fatal CHD), we assigned the next state as CHDW. Similarly, we assigned the next state as heart failure for incident fatal heart failure. For participants entering CHD and heart failure states at the same time (i.e., incident CHD combined with heart failure), we assigned the next state as CHD.

We applied our non-Markov framework to examine the course of heart disease. In particular, S_0_ to S_4_ states referred to healthy, at-metabolic risk, CHD, heart failure, and mortality states, respectively. By conditioning on past disease history, S_2|0_ and S_2|0_1_ referred to CHD without metabolic risk factors and CHD with metabolic risk factors, respectively, and S_3|0_, S_3|0_1_, S_3|0_2_, S_3|0_1_2_ referred to heart failure with no metabolic risk factors or CHD, heart failure only with metabolic risk factors, heart failure only with CHD, and heart failure with metabolic risk factors and CHD, respectively. We divided the data into subsets 1-4, starting from healthy, at-risk, CHD, and heart failure states, respectively. Within each subset, we changed the data into a long format by dividing them into small intervals by age and applied CSC models. We modeled age flexibly in cubic terms and included past disease history and the interaction terms with age as covariates in subsets 3 and 4. To estimate MALY, we used disability weights estimated from the Global Burden of Disease Study 2019,^23^ where the disability weights were 0.049, 0.072, and 0.074, for at-metabolic-risk (S_1|0_), CHD (S_2|0_), and heart failure (S_3|0_) states, respectively. For states with multimorbidity, a multiplicative approach was used to assign disability weights. For example, for CHD transited from healthy and at-metabolic-risk states, DW_2|0_1_ 1 − (1 − 0.049)(1 − 0.072).

### 3.3. Results

Our study included 15,027 participants at baseline, including 6831 men and 8196 women and 3917 Blacks and 11,110 Whites. Of the 15,027 participants, 4619 participants were healthy, and 10,408 participants were at metabolic risk at baseline. During a median of 27 years follow up, 2057 participants developed incidence of CHD, 3283 participants developed incidence of heart failure, and 7677 participants died. **Figure 3** shows the number of participants transited through the disease course over the follow-up period. The participants’ transition between states by sex and race is shown in **Figures S1-S4**.

**Figure 3.**
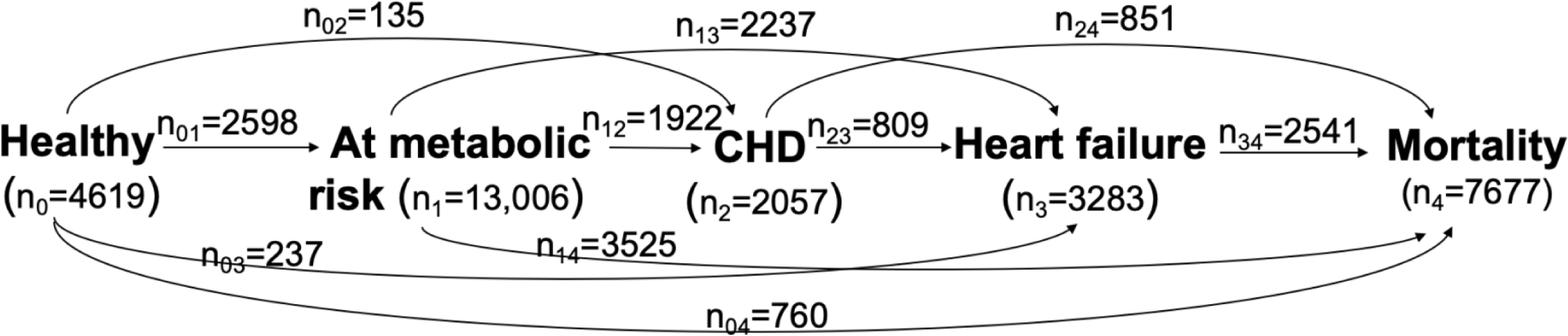
Participants’ transition through the disease course in the Atherosclerosis Risk in Communities Study (ARIC) study from 1987-2019 (n=15027).

**Table 1** shows the projected expected life years spent in each disease state. From the start of follow up, the expected life years were longest in at-metabolic-risk state (median: 18.25 years), followed by healthy state (median: 12.26 years). By summing up all disease states, the total expected life years was 25.69 (95% CI: 18.02, 34.16) years, which led to an estimated life expectancy of 79.87 (95% CI: 78.56, 82.06) years. By assigning a disability weight to account for multimorbidity in each state, the estimated MALY was 24.13 (95% CI: 16.55, 32.06) years, and estimated multimorbidity-adjusted life expectancy was 78.23 (95% CI: 75.95, 80.96) years. The multimorbidity-adjusted life expectancy was higher among women (79.29 [95% CI: 77.42, 81.57] years) than men (77.07 [95% CI: 74.37, 80.05] years) (**Table 2**), and was higher among Whites (79.04 [95% CI: 77.05, 81.31] years) than Blacks (75.49 [95% CI: 73.19, 79.30] years) (**Table 3**).

**Table 1.** Estimated disease state-specific life year and multimorbidity-adjusted life year (MALY) in the Atherosclerosis Risk in Communities Study (ARIC) study starting from 1987 (n=15027).

|  | **Disease states** | **Life Year (95% CI)** |
| --- | --- | --- |
| Disease-specific life year | Healthy (S<sub>0</sub>) | 12.26 (10.92, 15.67) |
|  | At metabolic risk (S<sub>1</sub>) | 18.25 (5.88, 29.48) |
|  | CHD without metabolic risk factors (S<sub>2\|0</sub>) | 0.28 (0.26, 0.31) |
|  | CHD with metabolic risk factors (S<sub>2\|0_1</sub>) | 1.41 (0.46, 1.87) |
|  | Heart failure with no metabolic risk factors or CHD (S<sub>3\|0</sub>) | 0.48 (0.34, 1.10) |
|  | Heart failure only with metabolic risk factors (S<sub>3\|0_1</sub>) | 1.63 (0.76, 1.74) |
|  | Heart failure only with CHD (S<sub>3\|0_2</sub>) | 0.05 (0.04, 0.06) |
|  | Heart failure with metabolic risk factors and CHD (S<sub>3\|0_1_2</sub>) | 0.35 (0.11, 0.48) |
| Unweighted | Unweighted life year | 25.69 (18.02, 34.16) |
|  | Unweighted life expectancy | 79.87 (78.56, 82.06) |
| Multimorbidity-adjusted | Multimorbidity-adjusted life year (MALY) | 24.13 (16.55, 32.06) |
|  | Multimorbidity-adjusted life expectancy | 78.23 (75.95, 80.96) |

**Table 2.** Estimated disease state-specific life year and multimorbidity-adjusted life year (MALY) by sex i in the Atherosclerosis Risk in Communities Study (ARIC) study starting from 1987.

|  |  | Life Year (95% CI) |  |
| --- | --- | --- | --- |
|  |  | Women (n=8196) | Men (n=6831) |
| Disease-specific<br>life year | Healthy (S<sub>0</sub>) | 12.39 (11.07, 15.41) | 12.08 (10.78, 16.24) |
|  | At metabolic risk (S<sub>1</sub>) | 18.71 (6.58, 31.26) | 16.99 (5.10, 27.36) |
|  | CHD without metabolic risk factors (S<sub>2\|0</sub>) | 0.13 (0.10, 0.19) | 0.49 (0.36, 0.61) |
|  | CHD with metabolic risk factors (S<sub>2\|0_1</sub>) | 1.18 (0.46, 1.43) | 1.75 (0.48, 2.49) |
|  | Heart failure with no metabolic risk factors or CHD (S<sub>3\|0</sub>) | 0.41 (0.32, 0.87) | 0.44 (0.30, 0.94) |
|  | Heart failure only with metabolic risk factors (S<sub>3\|0_1</sub>) | 1.90 (0.90, 2.00) | 1.27 (0.51, 1.37) |
|  | Heart failure only with CHD (S<sub>3\|0_2</sub>) | 0.02 (0.01, 0.03) | 0.09 (0.06, 0.12) |
|  | Heart failure with metabolic risk factors and CHD (S<sub>3\|0_1_2</sub>) | 0.33 (0.12, 0.41) | 0.47 (0.13, 0.66) |
| Unweighted | Unweighted life year | 27.73 (19.03, 35.40) | 23.78 (16.93, 32.02) |
|  | Unweighted life expectancy | 81.04 (80.11, 83.02) | 78.61 (76.91, 81.01) |
| Multimorbidity-adjusted | Multimorbidity-adjusted life year (MALY) | 25.63 (17.51, 33.80) | 22.23 (15.52, 30.67) |
|  | Multimorbidity-adjusted life expectancy | 79.29 (77.42, 81.57) | 77.07 (74.37, 80.05) |

**Table 3.** Estimated disease state-specific life year and multimorbidity-adjusted life year (MALY) by race in the Atherosclerosis Risk in Communities Study (ARIC) study starting from 1987.

|  |  | Life Year (95% CI) |  |
| --- | --- | --- | --- |
|  |  | White (n=11,110) | Black (n=3917) |
| Disease-specific life year | Healthy (S<sub>0</sub>) | 12.55 (11.23, 16.11) | 11.11 (9.23, 14.12) |
|  | At metabolic risk (S<sub>1</sub>) | 18.18 (6.01, 29.85) | 17.88 (6.54, 26.74) |
|  | CHD without metabolic risk factors (S<sub>2\|0</sub>) | 0.32 (0.28, 0.35) | 0.11 (0.07, 0.18) |
|  | CHD with metabolic risk factors (S<sub>2\|0_1</sub>) | 1.41 (0.46, 1.98) | 1.25 (0.46, 1.51) |
|  | Heart failure with no metabolic risk factors or CHD (S<sub>3\|0</sub>) | 0.40 (0.30, 0.82) | 0.36 (0.26, 0.74) |
|  | Heart failure only with metabolic risk factors (S<sub>3\|0_1</sub>) | 1.55 (0.69, 1.66) | 1.88 (0.93, 1.97) |
|  | Heart failure only with CHD (S<sub>3\|0_2</sub>) | 0.07 (0.06, 0.09) | 0.04 (0.02, 0.06) |
|  | Heart failure with metabolic risk factors and CHD (S<sub>3\|0_1_2</sub>) | 0.32 (0.10, 0.46) | 0.38 (0.13, 0.48) |
| Unweighted | Unweighted life year | 26.47 (18.44, 34.82) | 24.12 (16.62, 30.81) |
|  | Unweighted life expectancy | 80.67 (79.72, 82.48) | 77.20 (75.60, 80.69) |
| Multimorbidity-adjusted | Multimorbidity-adjusted life year (MALY) | 24.43 (16.98, 32.87) | 22.22 (15.26, 29.27) |
|  | Multimorbidity-adjusted life expectancy | 79.04 (77.05, 81.31) | 75.49 (73.19, 79.30) |

For healthy participants at baseline, the most likely disease paths estimated from our framework are: “Healthy → at metabolic risk → mortality” (37%), “Healthy → mortality” (21%), “Healthy → at metabolic risk → heart failure → mortality” (19%), and “Healthy → at metabolic risk → CHD → mortality” (8%) (**Table 4**). For participants at metabolic risk at baseline, 51% of participants were estimated to transit through the path “At metabolic risk → mortality”, 28% of participants were through the path “At metabolic risk → heart failure → mortality”, and 13% of participants were through the path “At metabolic risk → CHD → mortality”. The estimated disease paths were similar between women and men (**Table 5**), and similar between Whites and Blacks (**Table 6**).

**Table 4.** Estimated disease path in the Atherosclerosis Risk in Communities Study (ARIC) study starting from 1987 (n=15027).

| **Disease path** | **Percentage (%)** |
| --- | --- |
| **Start from healthy state ($S_0$)** |  |
| Healthy → mortality ($S_0 \rightarrow S_4$) | 21 |
| Healthy → at metabolic risk → mortality ($S_0 \rightarrow S_1 \rightarrow S_4$) | 37 |
| Healthy → CHD → mortality ($S_0 \rightarrow S_2 \rightarrow S_4$) | 2 |
| Healthy → at metabolic risk → CHD → mortality ($S_0 \rightarrow S_1 \rightarrow S_2 \rightarrow S_4$) | 8 |
| Healthy → heart failure → mortality ($S_0 \rightarrow S_3 \rightarrow S_4$) | 5 |
| Healthy → at metabolic risk → heart failure → mortality ($S_0 \rightarrow S_1 \rightarrow S_3 \rightarrow S_4$) | 19 |
| Healthy → CHD → heart failure → mortality ($S_0 \rightarrow S_2 \rightarrow S_3 \rightarrow S_4$) | 2 |
| Healthy → at metabolic risk → CHD → heart failure → mortality ($S_0 \rightarrow S_1 \rightarrow S_2 \rightarrow S_3 \rightarrow S_4$) | 5 |
| **Start from at-metabolic-risk state ($S_1$)** |  |
| At metabolic risk → mortality ($S_1 \rightarrow S_4$) | 51 |
| At metabolic risk → CHD → mortality ($S_1 \rightarrow S_2 \rightarrow S_4$) | 13 |
| At metabolic risk → heart failure → mortality ($S_1 \rightarrow S_3 \rightarrow S_4$) | 28 |
| At metabolic risk → CHD → heart failure → mortality ($S_1 \rightarrow S_2 \rightarrow S_3 \rightarrow S_4$) | 8 |

**Table 5.** Estimated disease path by sex in the Atherosclerosis Risk in Communities Study (ARIC) study starting from 1987.

| Disease path | Percentage (%) |  |
| --- | --- | --- |
|  | Women<br>(n=8196) | Men<br>(n=6831) |
| Start from healthy state ($S_0$) | | |
| Healthy → mortality ($S_0 \rightarrow S_4$) | 18 | 26 |
| Healthy → at metabolic risk → mortality ($S_0 \rightarrow S_1 \rightarrow S_4$) | 41 | 31 |
| Healthy → CHD → mortality ($S_0 \rightarrow S_2 \rightarrow S_4$) | 2 | 4 |
| Healthy → at metabolic risk → CHD → mortality<br>($S_0 \rightarrow S_1 \rightarrow S_2 \rightarrow S_4$) | 7 | 9 |
| Healthy → heart failure → mortality ($S_0 \rightarrow S_3 \rightarrow S_4$) | 6 | 5 |
| Healthy → at metabolic risk → heart failure → mortality<br>($S_0 \rightarrow S_1 \rightarrow S_3 \rightarrow S_4$) | 21 | 17 |
| Healthy → CHD → heart failure → mortality<br>($S_0 \rightarrow S_2 \rightarrow S_3 \rightarrow S_4$) | 1 | 3 |
| Healthy → at metabolic risk → CHD → heart failure → mortality<br>($S_0 \rightarrow S_1 \rightarrow S_2 \rightarrow S_3 \rightarrow S_4$) | 4 | 5 |
| Start from at-metabolic-risk state ($S_1$) | | |
| At metabolic risk → mortality ($S_1 \rightarrow S_4$) | 55 | 48 |
| At metabolic risk → CHD → mortality ($S_1 \rightarrow S_2 \rightarrow S_4$) | 10 | 16 |
| At metabolic risk → heart failure → mortality ($S_1 \rightarrow S_3 \rightarrow S_4$) | 30 | 27 |
| At metabolic risk → CHD → heart failure → mortality<br>($S_1 \rightarrow S_2 \rightarrow S_3 \rightarrow S_4$) | 6 | 9 |

**Table 6.** Estimated disease path by race in the Atherosclerosis Risk in Communities Study (ARIC) study starting from 1987.

| Disease path | Percentage (%) |  |
| --- | --- | --- |
|  | White<br>(n=11,110) | Black<br>(n=3917) |
| Start from healthy state ($S_0$) | | |
| Healthy → mortality ($S_0 \rightarrow S_4$) | 21 | 22 |
| Healthy → at metabolic risk → mortality ($S_0 \rightarrow S_1 \rightarrow S_4$) | 37 | 36 |
| Healthy → CHD → mortality ($S_0 \rightarrow S_2 \rightarrow S_4$) | 3 | 1 |
| Healthy → at metabolic risk → CHD → mortality<br>($S_0 \rightarrow S_1 \rightarrow S_2 \rightarrow S_4$) | 9 | 7 |
| Healthy → heart failure → mortality ($S_0 \rightarrow S_3 \rightarrow S_4$) | 6 | 6 |
| Healthy → at metabolic risk → heart failure → mortality<br>($S_0 \rightarrow S_1 \rightarrow S_3 \rightarrow S_4$) | 18 | 22 |
| Healthy → CHD → heart failure → mortality<br>($S_0 \rightarrow S_2 \rightarrow S_3 \rightarrow S_4$) | 2 | 1 |
| Healthy → at metabolic risk → CHD → heart failure → mortality<br>($S_0 \rightarrow S_1 \rightarrow S_2 \rightarrow S_3 \rightarrow S_4$) | 5 | 4 |
| Start from at-metabolic-risk state ($S_1$) | | |
| At metabolic risk → mortality ($S_1 \rightarrow S_4$) | 52 | 50 |
| At metabolic risk → CHD → mortality ($S_1 \rightarrow S_2 \rightarrow S_4$) | 14 | 11 |
| At metabolic risk → heart failure → mortality ($S_1 \rightarrow S_3 \rightarrow S_4$) | 26 | 32 |
| At metabolic risk → CHD → heart failure → mortality<br>($S_1 \rightarrow S_2 \rightarrow S_3 \rightarrow S_4$) | 8 | 7 |

## 4. DISCUSSION

In this study, we derived two summary estimates to characterize disease course, MALY which accounts for multimorbidity and estimates the adjusted life year in full health, and disease path which estimates path of state transition over disease course. We used the two estimates to characterize the course of heart disease and showed that they have potential to be used in chronic disease prevention.

From a methodology perspective, the summary estimates derived from our study may advance multi-state modeling in disease prevention. Multi-state Markov models have been used to investigate course of chronic disease, which estimate hazard ratio of exposures with disease state transition.^1, 2^ Semi-Markov model models relax the Markov assumption by allowing future states to depend on duration in current state.^24, 25^ However, these models may be inappropriate to model the course of chronic disease, as past states of chronic disease can interplay in affecting future states which violates the Markov assumption. Most importantly, transition probabilities estimated from these models are time-specific and are not straightforward for public health interpretation. Our non-Markov framework conditions on past states to divide disease states into substates to convert a non-Markov to Markov process. As the substates memorize past disease history and inform multimorbidity, the transition rates between substates can be used to generate MALY and disease path. These summary estimates characterize the whole disease course, are easy to interpret, can be used for Markov process (as a special case of non-Markov process), and will potentially advance multi-state modeling in comparative effectiveness of intervention, which compares effect of intervention to inform decision-making. Focusing on one disease endpoint may not allow to fully assess benefits and harms of intervention, which leads to inaccurate estimation of intervention effect and misidentify optimal intervention. MALY estimates life year across the disease course accounting for multimorbidity and can be used by health policy makers to better evaluate intervention effects to facilitate policy decision-making. Disease paths characterize long-term disease course and can facilitate medical resources planning for effective early prevention of disease progression.

Heart disease has been the leading cause of mortality in the U.S.,^26^ and imposes a heavy economic burden to families and society.^27^ Previous epidemiological studies have generated fruitful knowledge about heart disease prevention,^28–32^ however, most of them have focused on one single endpoint, such as hypertension, CHD, or heart failure, or a composite endpoint such as CVD.^21, 33–36^ However, the development of heart disease is a multi-state process that is biologically inseparable: Hypertension and hyperlipidemia alter vascular elasticity, narrow vascular lumen, increase cardiac work, and result in heart disease.^37, 38^ Although a few studies applied multi-Markov models to investigate heart disease prevention,^39, 40^ the inconsistent associations of exposures with state transitions make it difficult to draw an conclusion. For example, the China Kadoorie Biobank showed a healthy lifestyle played different roles in states transitions, which was associated with a lower rate of transition of healthy to cardiometabolic disease (CMD) but not associated with transition from CMD to mortality.^39^ The MALY derived from our study synthesizes transition rates across disease states into one estimate, which can be used to evaluate the effects of prevention across disease course.

Our study showed that the longest life year was from at-metabolic-risk state, providing evidence to formulate prevention strategies specifically targeting at-metabolic-risk population. The American Heart Association “Life’s Essential 8” should be adopted at early states to prevent the development of heart disease, which includes maintaining a normal weight, consuming a healthy diet, being physically active, no smoking, and adequate amount of sleep.^41^ The life expectancy in the U.S. has drastically increased over the past 50 years, primarily driven by a continuous increase in survival rate after CVD prognosis.^42, 43^ The estimated life year for states with CHD or heart failure allows to estimate the demand for health services and the costs for treatment more precisely. Moreover, the estimated life year of disease states with CHD or heart failure was short and suggested the importance of secondary prevention of heart disease. For example, adopting a healthy lifestyle together with statin use have shown long-term benefits for secondary prevention of heart disease and mortality.^44, 45^

The estimated path of heart disease is of potential public health significance in the early prevention of heart disease. Our study found that the most likely path of heart disease progression was “healthy → at metabolic risk → mortality” followed by “Healthy → at metabolic risk → heart failure → mortality”. This highlights the importance of early screening of heart failure among participants with metabolic risk factors. Heart failure is commonly considered a consequence of CHD, where CHD can damage heart muscle and cause left ventricular dysfunction and heart failure.^46^ However, the incidence of heart failure has substantially increased in the past decades, with etiologies differing from CHD.^47^ To identify heart failure at early stage, heart failure-specific biomarker, B-type natriuretic peptide (BNP), can be used to identify heart failure before structural and functional changes of heart muscle become obvious.^48–51^ Moreover, echocardiogram-based predictors, such as left ventricular ejection fraction, can detect left ventricular dysfunction and heart muscle damage.^52^

Our study found that the estimated MALY was higher among women than men and higher among Whites than Blacks. Consistently, previous studies have shown that biological race and sex influence risk of heart disease:^53–57^ The prevalence of heart disease is higher in men than women,^54^ and higher among Blacks than Whites.^53^ One reason for this is due to the different prevalence of healthy lifestyles. The prevalence of obesity and being physically inactivity is higher among Blacks than Whites, although the prevalence of smoking is higher among Whites.^53^ Men are more likely to smoke, although they are more active than women.^55–57^ The other reason may be due to the racial disparities in accessing healthcare for heart disease prevention and treatment. Studies have shown that the overall health care use was higher among Whites than Blacks, and this have persisted for 6 decades and widened in recent years.^58, 59^

In conclusion, we proposed two summary estimates derived from a multi-state non-Markov framework and applied it to characterize the course of heart disease. The two estimates describe disease course in a summary manner and have potential use in chronic disease prevention.

## Data Availability

The data can be requested through Biologic Specimen and Data Repository Information Coordinating Center (BioLINCC).

---

**Figure S1.**
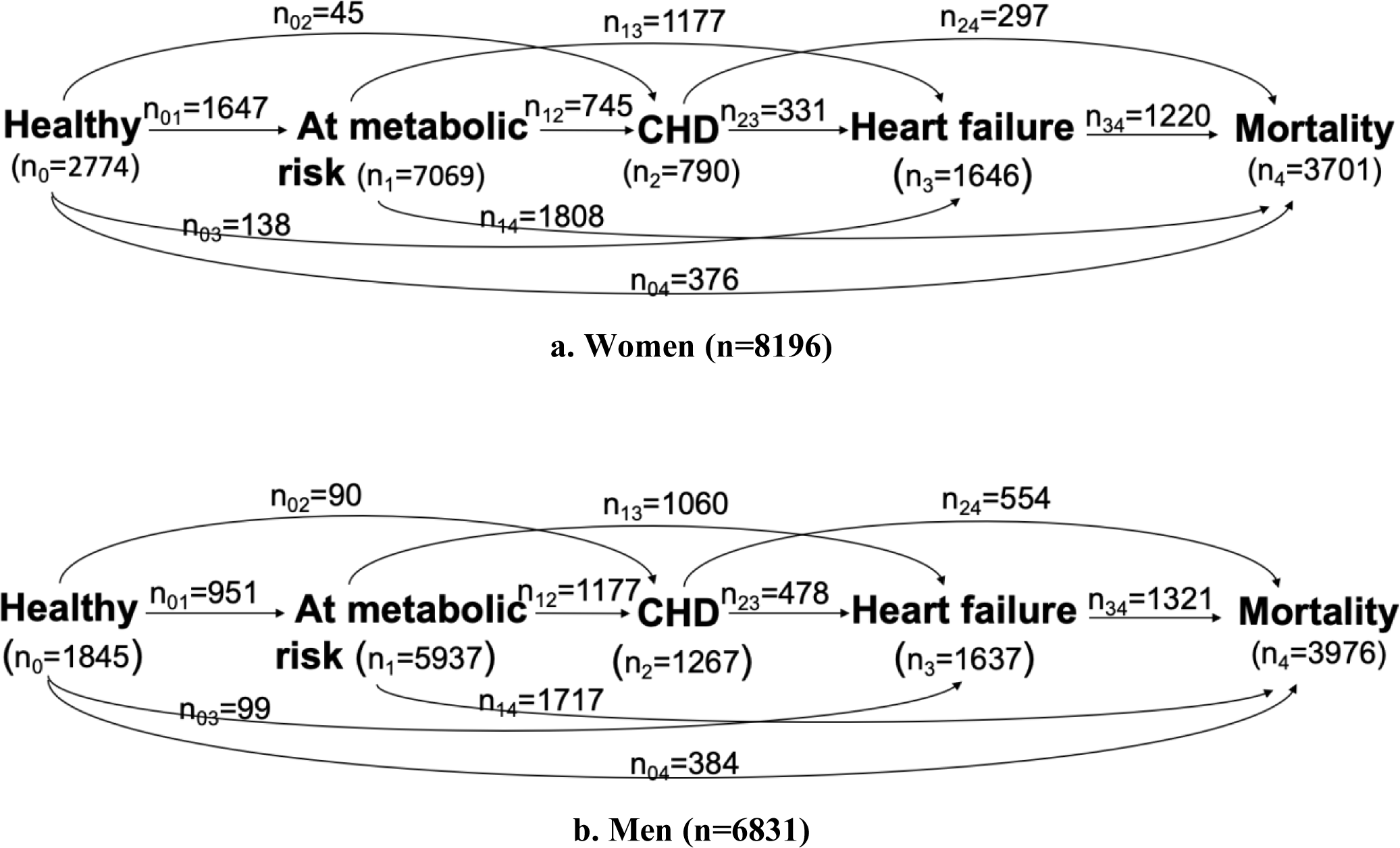
Participants’ transition through the disease course by sex in the Atherosclerosis Risk in Communities Study (ARIC) study starting from 1987.

**Figure S2.**
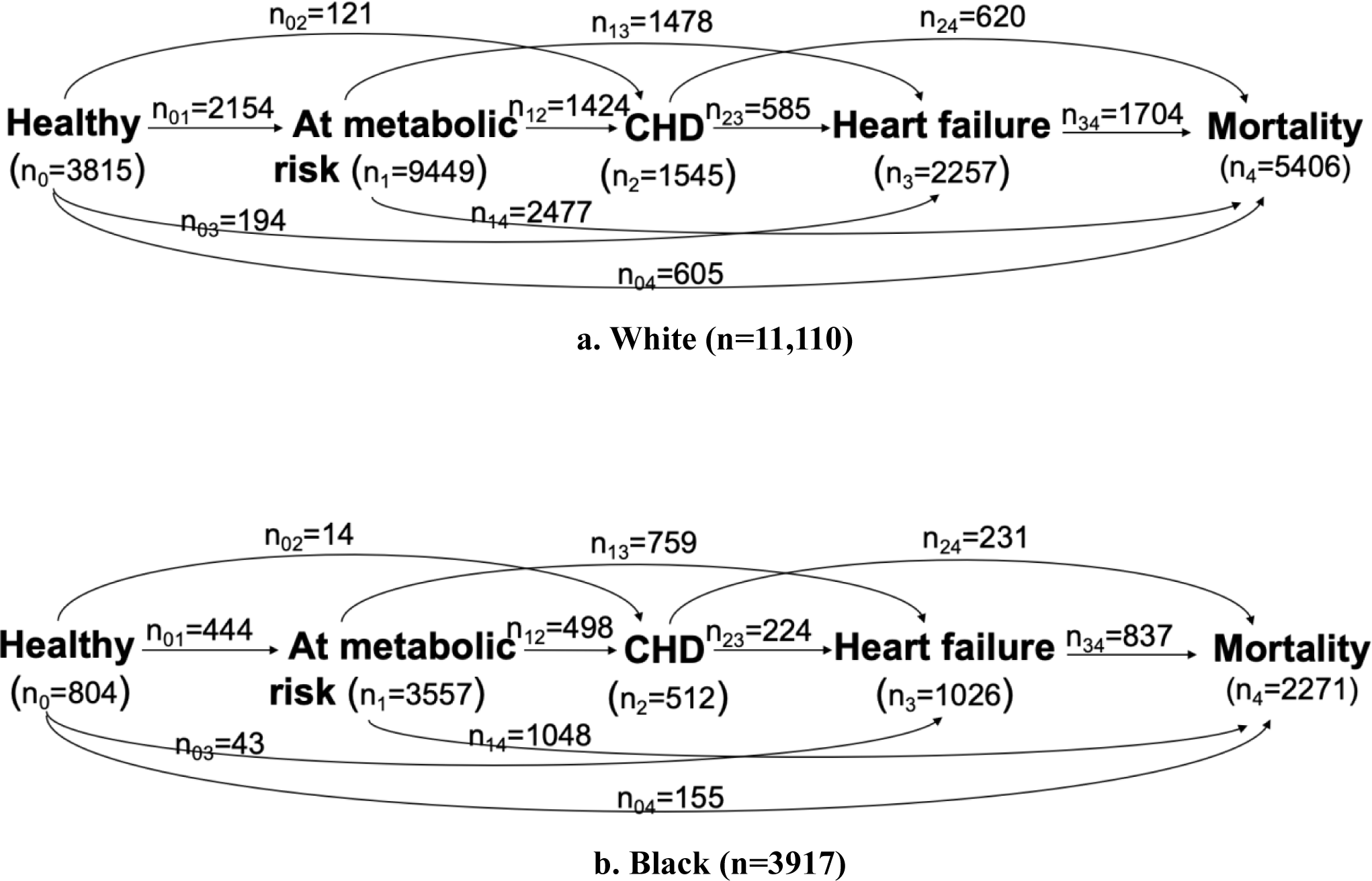
Participants’ transition through the disease course by race in the Atherosclerosis Risk in Communities Study (ARIC) study starting from 1987.

